# Predicting Long-term Evolution of COVID-19 by On-going Data using Bayesian Susceptible-Infected-Removed Model

**DOI:** 10.1101/2020.05.08.20094953

**Authors:** Shohei Hidaka, Takuma Torii

## Abstract

In this study, we propose a novel statistical method to predict a long-term epidemic evolution based on a on-going data. We developed a Bayesian framework for the Susceptible-Infected-Removed model (Bayesian SIR), and estimated its underlying parameters based on day-by-day timeseries of the cumulative number of infectious individuals. The new Baysian framework extends the deterministic SIR model to a probabilistic form, which provides an accurate estimation of the underlying system by a short and noisy data. We applied it to the data reported on the Coronavirus Disease 2019 (COVID-19), and made a month long prediction on its evolution. Our simulated test using past timeseries to predict the current data gives a reasonable reliablity of the proposed method. Our analysis of the current data detected and warned a rising trend in the countries in Central Asia, Middle East, and South America, while United States or European countries, which have already experienced large numbers of infected cases, are predicted to slow down in the increase.

## 1 Background and aims

The pandemic of the Coronavirus Disease 2019 (COVID-19) has been an unprecedent disaster and threat all over the world, since its first report in Wuhan, China in December 2019. Many countries in Asia, Europe, and North America have already experienced sharp increases in the number of infectious cases and deaths caused by COVID-19, and some of them observed slow down in the increase of the infectious cases and deaths. Several of the authorities of these countries have resorted lockdowns of major cities or whole countries, and stopped major body of social activities except for the minimum necessary one to keep citizens’ daily life. It is hypothetically effective to reduce person-to-person interaction in order to lower the basic production number, how many additional infected people on average each infected person causes, and reduce the outbreak size in each country. A powerful social resort such as lockdown, however, also causes a severe economical damage in society, which may also cause other types of social and economic crises. Thus, in this current situation, it is critical to estimate or predict the final or a long-term order of outbreak size, in order for the authority make to balance two distinct types of risks caused by COVID-19 and potential economic depression.

In this present study, we aim to develop a statistical model to predict the evolution of the infectious population size in a middle-term (month order) time scale using a given reported timeseries of cumulative number of infectious cases. It has been considered difficult to accurately predict the final outbreak size in general [Drake, 2005]. Perhaps, one of reason for this problem is that we still miss a proper statistical re-formulation of a theoretical model such as the Susceptible-Infected-Removed (SIR) epidemic model [Kermack and McKendrick, 1927], which is, in the original form, not appropriate to directly apply to a noisy data subject to non-infectious/ social impact taken by the human society.

There are a number of past attempts to give a quantitative prediction for the cumulative number of infectious cases. Some attempts employed a curve-fitting of theoretically assumed function such as logistic function [Batista, 2020, Fokas et al., 2020], others employed a statistical model designed to capture an epidemic dynamics, and provided a model-based analysis of the data [Andersson and Britton, 2012, Clancy et al., 2008]. The former type of approach often takes a more generic curve-fitting method, but has little theoretical justification for the fitted function, which the latter has. In contrast, the latter type of approach has theoretical rational and epidemiological background behind the model, but it is often too technically restricted to apply an empirical, noisy and unknown type of disease.

In this study, we developed a statistical modification of the deterministic SIR (DSIR) model [Kermack and McKendrick, 1927], which we call *Bayesian SIR* model (BSIR). BSIR is a probabilistic and generative model in which the individuals in the Susceptible, Infected, and Removed state stochastically transit one state to the other, as modeled in DSIR. In this sense, BSIR employed the theory-driven approach [Andersson and Britton, 2012, Clancy et al., 2008], while it is also data-driven to readily apply to a noisy empirical data. BSIR is essentially a probabilisstic, discrete-time and discrete-variable modification of DSIR, and reformulate the original differential equations to a difference equations. With this reformulation, we discovered a theretical relationship between an partially observed timeseries and the latent system parameters in BSIR.

## 2 SIR Model and its extension

The SIR model [Kermack and McKendrick, 1927] is one of the earliest models in the form of a set of differential equations as follows, which captures irreversible transition of population, from/to the three types of population, *Susceptible* (*S*), *Infected (I*), and *Recovered* (*R*), *S* + *I* + *R* = *N*.

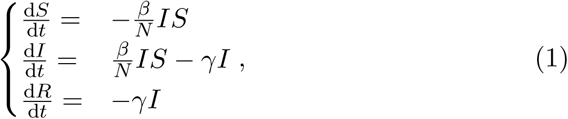

In a typical initial setting, vast majority of the whole population of interest is at Susceptible to the disease, and introduces a small portion of infected population in it (i.e., large *S* and small *I*). In the form of differential equation, the increase rate in the infected population is characterized by the proportion of the infected population to the whole population, and thus 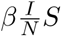 of people move from susceptible to infected state at every unit time with a certain rate constant *β*. Some portion of the infected population is recovered (sometime *removed* is more preferred as “death” also count as “recovered”) and immune from the infected population by *γI* with a certain rate *γ* every unit time. Thus, the standard SIR model has three parameters dominating the dynamics, the (initial susceptible) population size *N*, the increasing rate of recovery *γ*, and the increasing rate of infectious population size *β*.

There are many past theoretical and technical attempts to model various types of epidemic and developed statistical analysis for them. Andersson and Britton [Andersson and Britton, 2012] covered basic stochastic models such as the Reed-Frost model [Abbey, 1952], SIR model [Kermack and McKendrick, 1927], and their extension and variations, as well as their statistical analysis for a partially observed data. However, most of these models are formulated as an analogue of the original differential equations, in a form of continuous-time stochastic process [Andersson and Britton, 2012], and they are often technically difficult to adopt for increasing timeseries with unknown initial and terminal condition (but some may have a closed-form estimator [Clancy et al., 2008]).

The philosophy behind these existing theoretical work is more or less “post-epidemic analysis” with near-complete data and findings on well-known epidemic, rather than practical analysis of on-going data to predict evolution of an unknown type of epidemic. That is, the major body of theoretical analysis is dedicated for analysis of vaccination efficacy and proposal for the vaccination policy, after an epidemic has been experienced fully (see also Chapter 12 of [Andersson and Britton, 2012]). For instance, Clancy and O’Neill [Clancy et al., 2008] have derived a closed-form estimator of the basic production number for a given complete data following a stochastic SIR model. This is of course quite important by its own right, in order to prevent the next epidemic and reduce causalities. It is, however, less useful, when we *are* going through little known epidemic and had no vaccine against it for time being.

Given this background of the related theoretical literature, the new model is designed to estimate the latent nature of outbreak such as the basic reproduction number and the initial susceptible population size, which are hypothetically assumed in the DSIR model, using a short and on-going timeseries of the cumulative number of infectious individuals. Although our probabilistic model is inspired by DSIR model in the philosophy level, its computatoinal architecture is quite different from the existing ones. First, many of the existing statistical SIR models is formulated in continuous-time (but not all), but our model is discrete time, which is also considered a sophistication of the Reed-Frost “chain of binoamial” model [Abbey, 1952], in order to incorporate empirical data typically reported daily. Second, the new model is designed to produce a relatively easily computable estimator, rather than being faithful to the original SIR model. Thirdly, the model is fully data-driven – neither system parameters nor latent variables need to be pre-specified, but all of governing parameters are estimated from partially observed data. These three design principles are in our mind, we chose Bayesian statistical framework, in which the observed and latent infected population size and SIR-like characteristic parameters, as random variables, are analyzed over discrete time.

Comparable with the differential equation (1) of the state variables *(S(t),I(t), R(t))* ∊ *ℝ*^3^, Baysian SIR model has the three discrete state variables *(S_t_, I_t_, R_t_)* ∊ ℤ^3^ over discrete time *t* = 0,1,…. The Baysian SIR model is designed to capture the essential nature of the differential equation (1) by the difference equations for any *t* = 1, 2,…

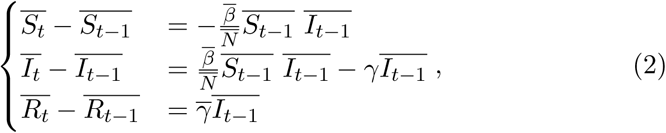

where each variable 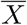 indicates the expected value of the random variable *X*.

As an empirical dataset of the on-going epidemic evolution, we do not know the system parameters *N, β, γ* as well as *S_t_, I_t_, R_t_* exactly, but we can only access (sub-sample of) the cumulative number of infected individuals *F_t_* = *N − S_t_*. Suppose that we are given a g timeseries of the cumulative number of infectious cases *F_t_* at day *t* = 0,1,…,*T*

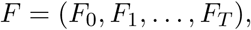

and BSIR predicts the continued hypothetical time series up to a specified time 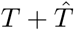 in future

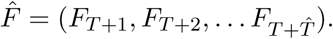

An input timeseries *F* is typically very short (≈ 30 by discarding early fluctuated timeseries) for make any generic method to give a sufficiently accurate prediction. As an example, Figure 1 shows a generated timeseries of *F*(*t*) of DSIR and *F_t_* of BSIR with the same parameter *N* = 10^6^,*β* = 0.5,*γ* = 0.2. These two models produce quite similar timeseries, although their time constant or lag may be slightly different between them due to randomness due to early stage of the development in BSIR.

**Figure 1:**
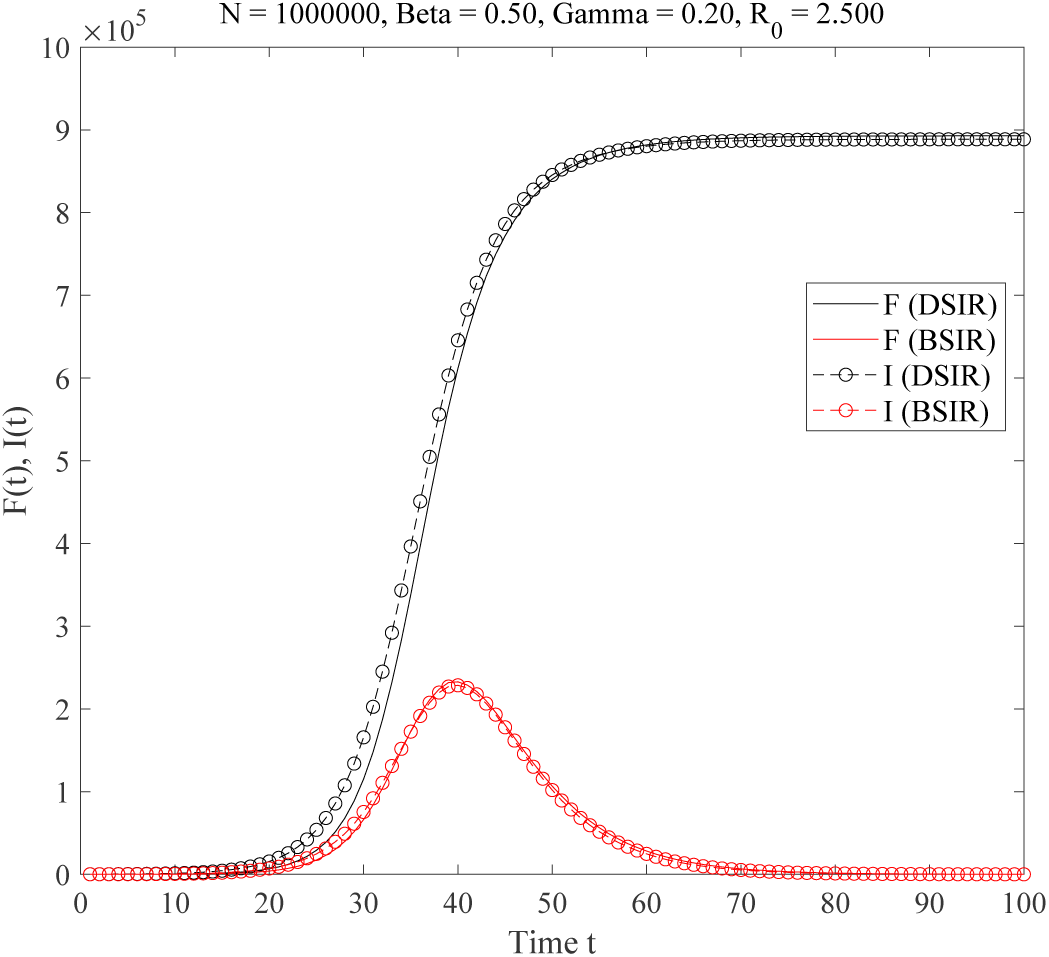
The cumulative number of infected individuals *F*(*t*) = *N* − *S*(*t*) and the number of infected individuals *I*(*t*) in DSIR and BSIR model.

It seems challenging to find all parameters only with this timeseries alone by existing techniques, since it seems insufficient to decide SIR model with two independent variables in the three state variable *S, I, R*. In BSIR, the timeseries of cumulative number of infected individuals *F* gives a crucial statistics to estimate the parameter *N, β, γ*, as the expected values of these random variables always hold the identity for any *t* = 2, 3, ... (see also Lemma 1 in the Supplement)

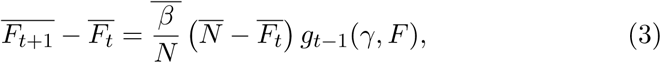

where 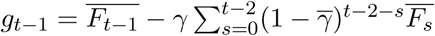.

Identity (3) motivates us to plot this relationship between timeseries *F*, which we call *cumulative-to-difference* plot (CD plot), to characterize a BSIR by the series of points 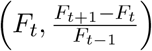 on the plane ℝ^2^. Figure 2 shows CD plot. In the CD plot, there is an asymptotic line drawn by letting *g_t_*_−1_ in (3) be constant, and it hits the latent cumulative number of infected individuals *N* on the X-axis in the limit *t → ∞*. Thus, it suggest that a naive way to predict the evolution of epidemic is to extrapolate an existing data to an asymptotic line on the CD plot. However, this simple method is not effective in practice – a noisy empirical timeseries does not allow to estimate the asymptotic line unless the sample size is large enough (see also Section **??** for the numrical simulation and more discussion on this point). Accordingly, we developed a Bayesian estimator of the parameters *N, β, γ* and other latent variables to extract full information underlying the series *F*.

**Figure 2:**
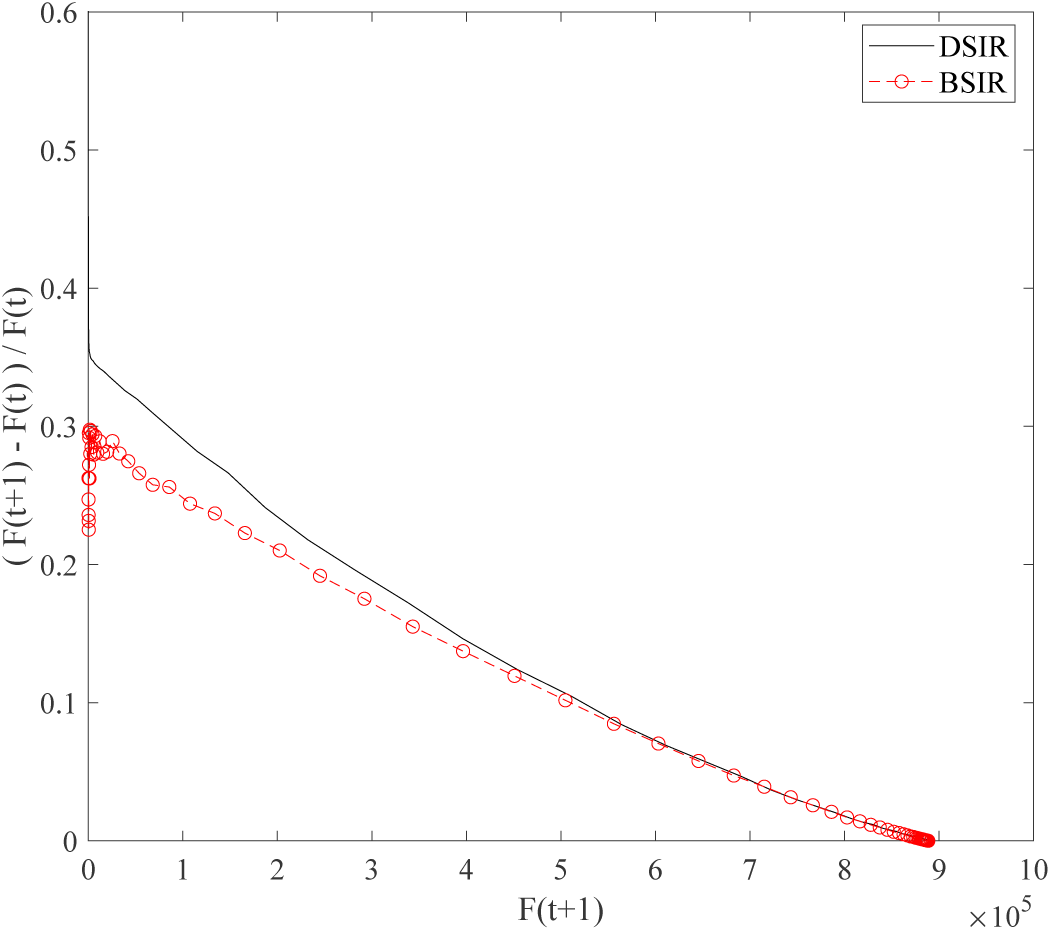
The increase ratio 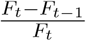 as a function of the cumulative number of infected individuals *F*(*t*) = *N − S(t)* in DSIR and BSIR model.

## 3 Prediction with empirical data

Therefore, on behave of just a simple linear-regression based approach, we incorporated a domain knowledge on epidemic mechanism, embodied as the DSIR model, to design BSIR applicable to the empirical report on daily timeseries of infectious cases (see also Section **??** for the detail of BSIR).

BSIR is a fully automated statistical analyzer and predictor of evolution of epidemic timeseries. We provided a series of analyses with both simulated and empirical data in Supplement (Section 4 and 5). This analyses suggest that our method gives a reliable prediction up to a month long future.

To demonstrate its effitiveness in predicting epidemic dynamics, we applied BSIR to the dataset of United States collected and published by Johns Hopkins University Center for System Science and Engineering [for Systems Science and at Johns Hopk accessed on April 27th, 2020 (data up to April 26th). These two countries at the present point have reported two largest cumulative number of infectious cases. To see the prediction accuracy, we applied BSIR to the data upto March 12th (1.5 month ago from the date of analysis), and every 14 days up to April 23rd. Figure 3 to 6 show the snapshots of prediction to these datasets. In Figure 3 to 6, the median prediction and the credible interval in dark color points and region, and the reported timeseries (but not used for prdiction) is in light color. The data used for prediction is in dark color. As the prediction with early date showed a large credible interval, the logarithm scale on Y axis was shown. The descriptive and predictive statistics are summarized in Table 1. On March 12th, 26th, April 9th, 23rd, and 26th, it has reported 1663, 83836, 462780, 869,170 and 965,785 cumulative number of infected individuals, respectively. As early as 1.5 month before over 900 hundreds count or 523 times of increase, BSIR can predict its potential risk by the median estimate 2,289,986 at the point of April 26th (965,785) and a large 95% credible interval from 23,905 to 86,808,211 (see also Section ?? for the detail and discussion about BSIR estimates). Two weeks later, BSIR made a generally consistent prediction, the median 1,237,030 (to the actual 965,785) with a smaller 95% credible interval from 637,004 to 55,303,162. Prediction made using the dataset up to April 9th underestimated it, but this may reflect a potential effect of lockdown, which started March 19th (Calfornia) and onward (other states started on the latest date March 24th). As the current model does not assume external effect on an epidemic dynamics, prediction may be affected by such social/ political action in the community.

**Figure 3:**
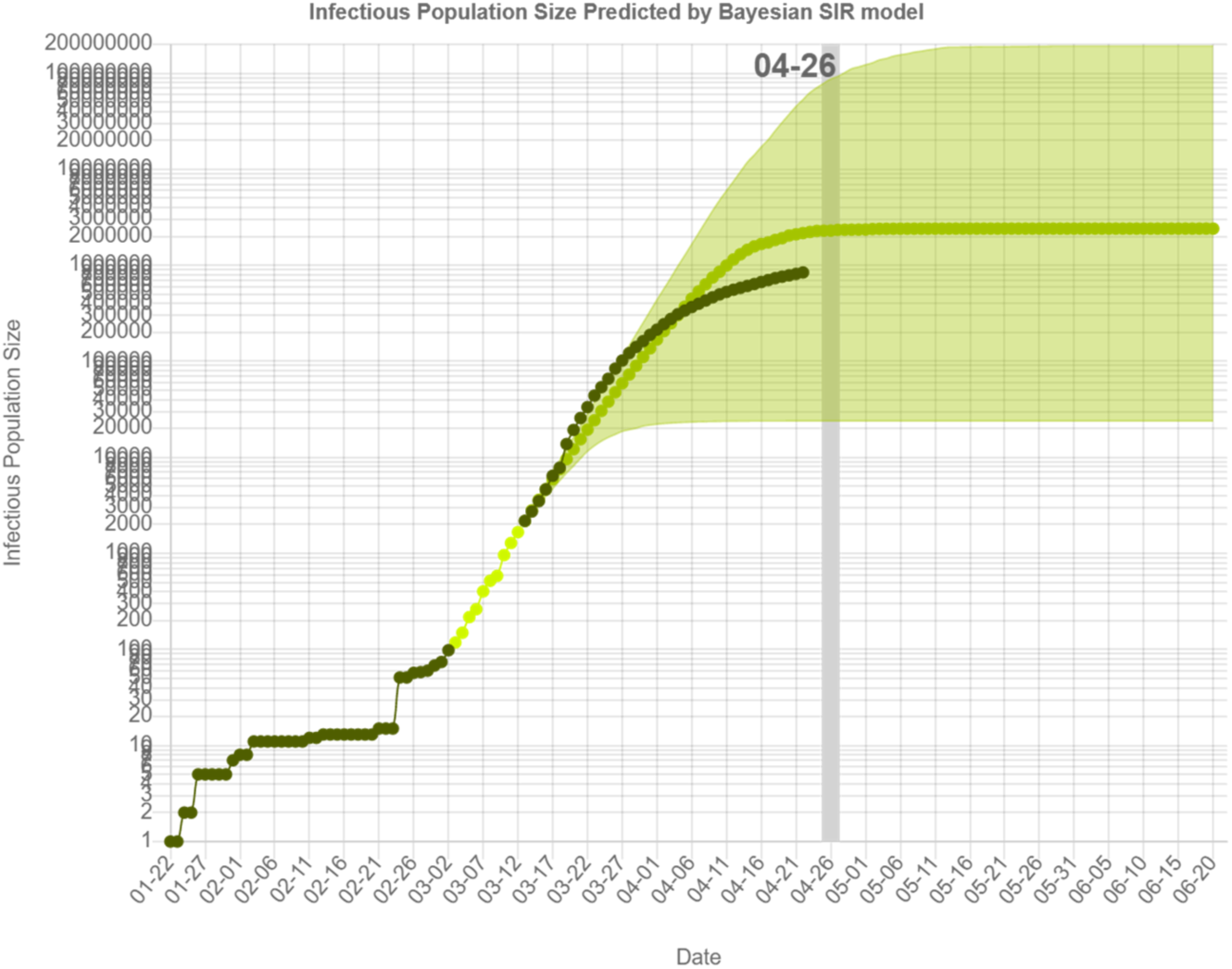
Prediction of the cumulative number of individuals in United States, estimated using the data up to March 12th.

**Figure 4:**
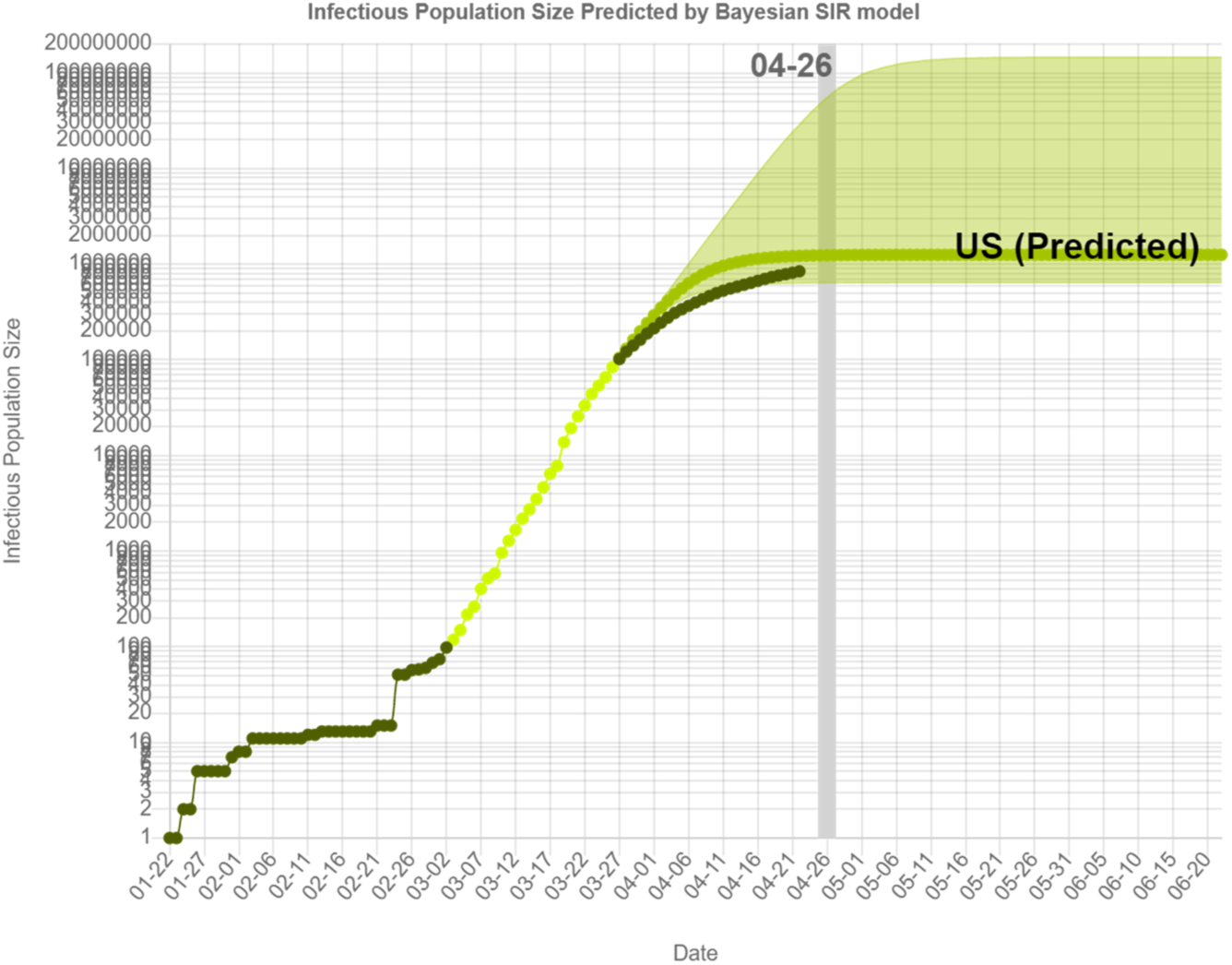
Prediction of the cumulative number of individuals in United States, estimated using the data up to March 26th.

**Figure 5:**
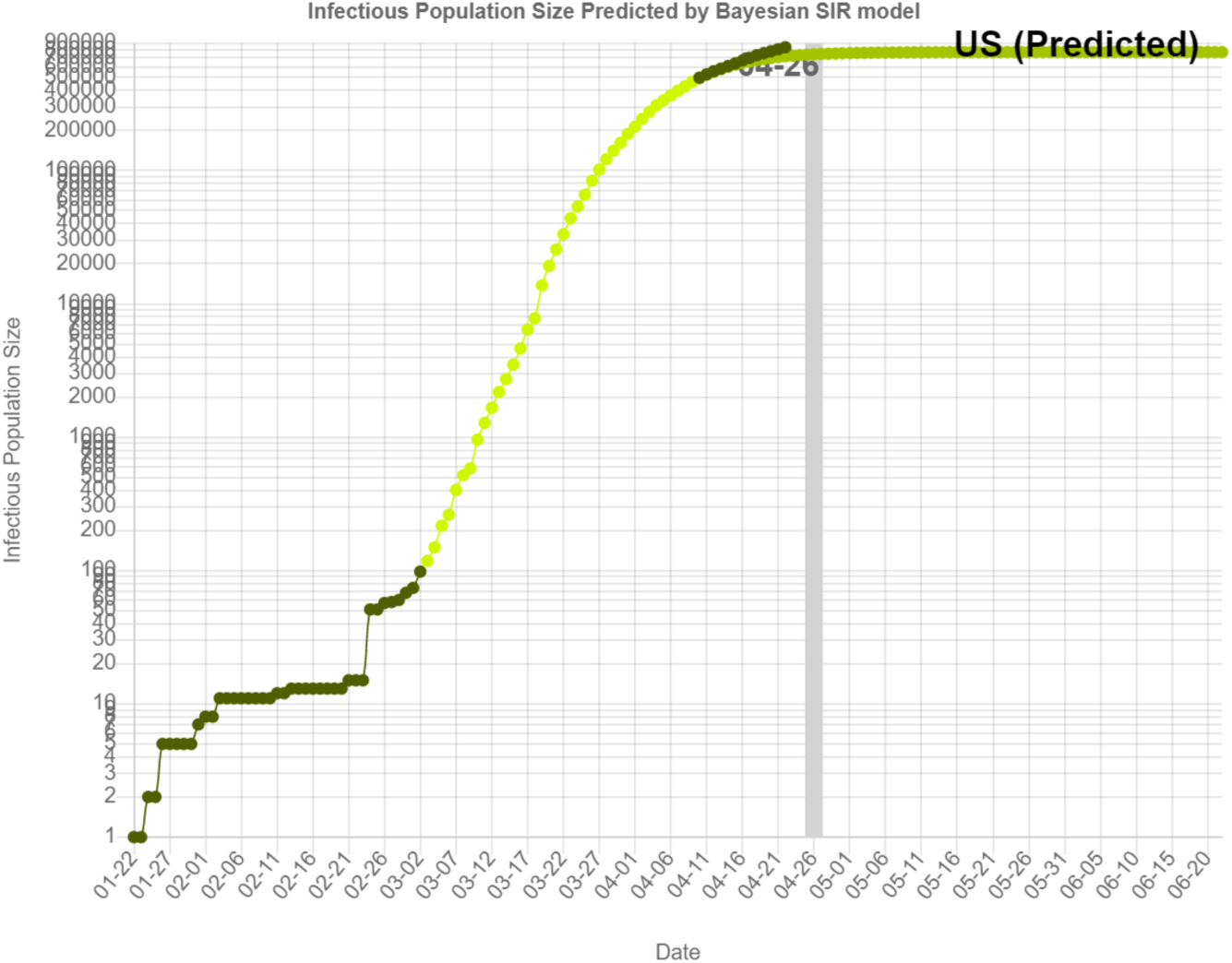
Prediction of the cumulative number of individuals in United States, estimated using the data up to April 9th.

**Figure 6:**
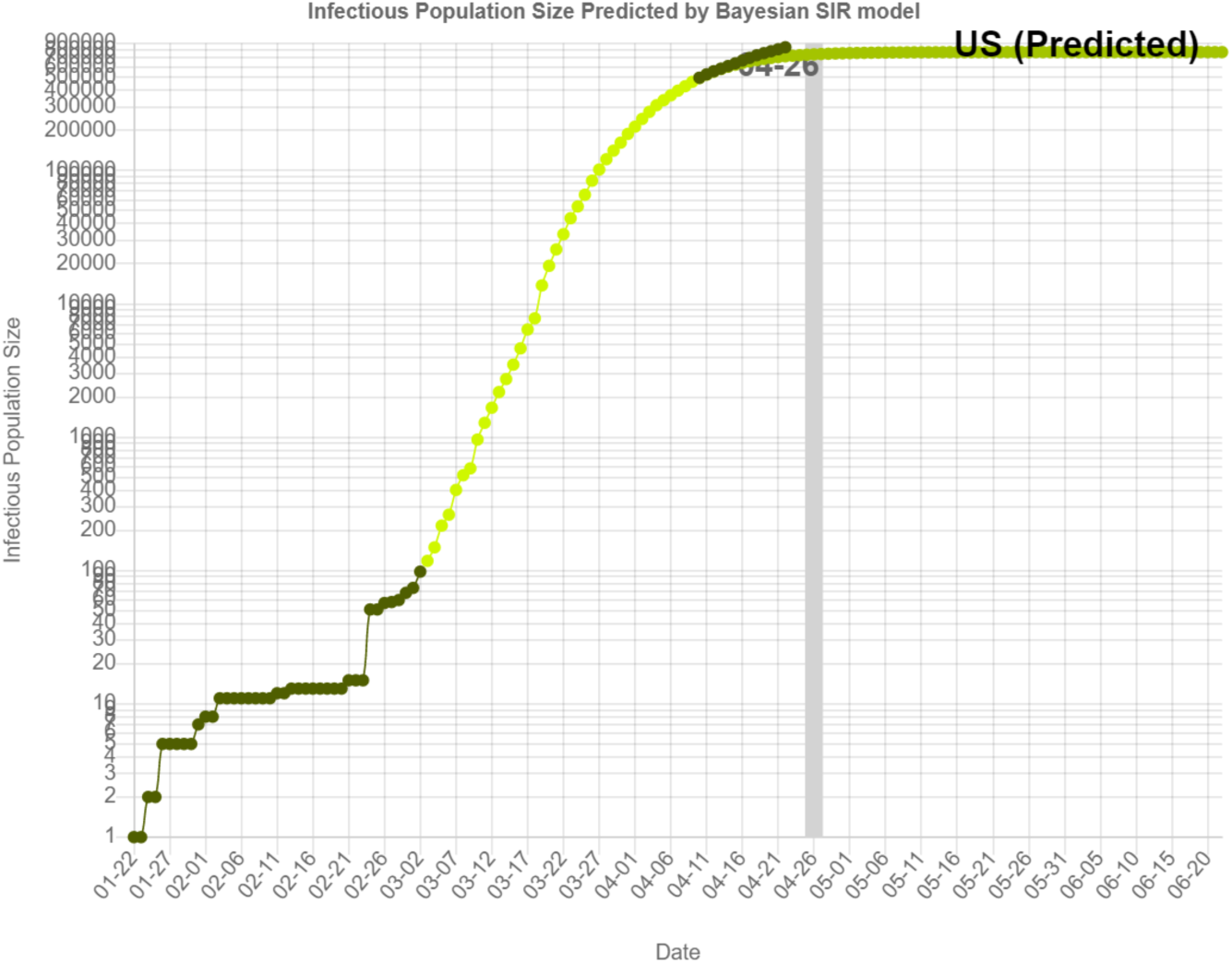
Prediction of the cumulative number of individuals in United States, estimated using the data up to April 23rd.

**Table 1:** Prediction of the cumulative number of infected cases in US on April 26th made by BSIR model using the past data (1.5 month ago and every 14 days).

| Data up to | Reported | Median | Credible Interval |
| --- | --- | --- | --- |
| Mar. 12th | 1,663 | 2,289,986 | (23905, 86808211) |
| Mar. 26th | 83,836 | 1,237,030 | (637004, 55303162) |
| Apr. 9th | 462,780 | 743634 | (732700, 756243) |
| Apr. 23rd | 869,170 | 933800 | (935749, 931797) |
| Apr. 26rd | 965,785 | - | - |
| Prediction on |  |  |  |
| Aug. 4th | - | 1,408,566 | (1383435, 1435755) |

The cumulative-difference plot with the data of US up to April 26th and prediction was shown in Figure 7. The empirical data points in the CD plot has a certain level of noise, but the BSIR captures a general trend of the empirical data and extrapolate to the estimated asymptotic line. Based on this point of prediction (data available on April 26th), the predicted cumulative number of infected individuals is 1,408,566 (the 95% credible interval is 1383435 ≤ *F*(*t*) ≤ 1435755) on August 4th, which is supposed the near final outbreak size, if the current socio-political status in US was kept.

**Figure 7:**
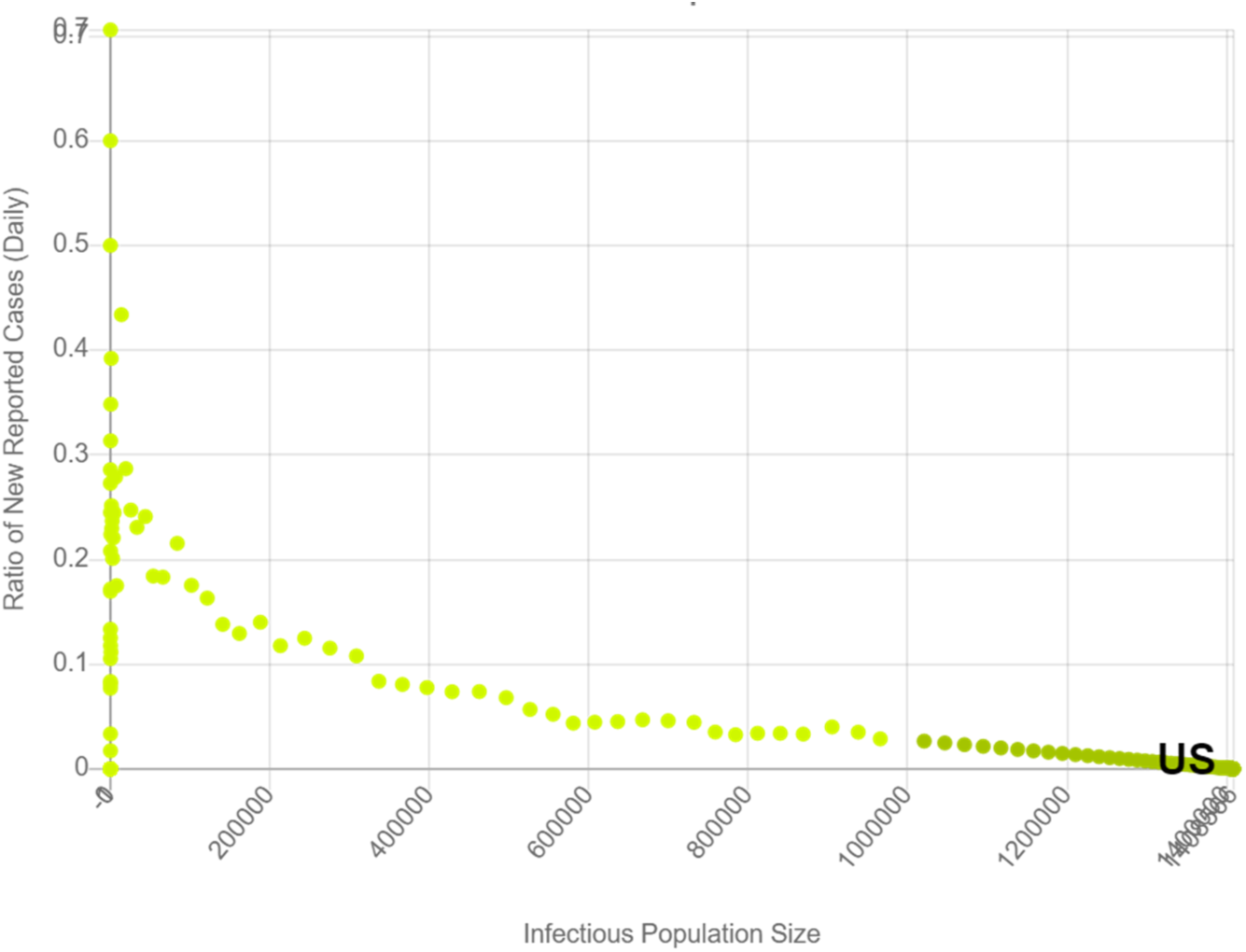
Prediction of the cumulative number of individuals in United States in the form of cumulative-difference plot, estimated using the data up to April 26th.

**Figure 8:**
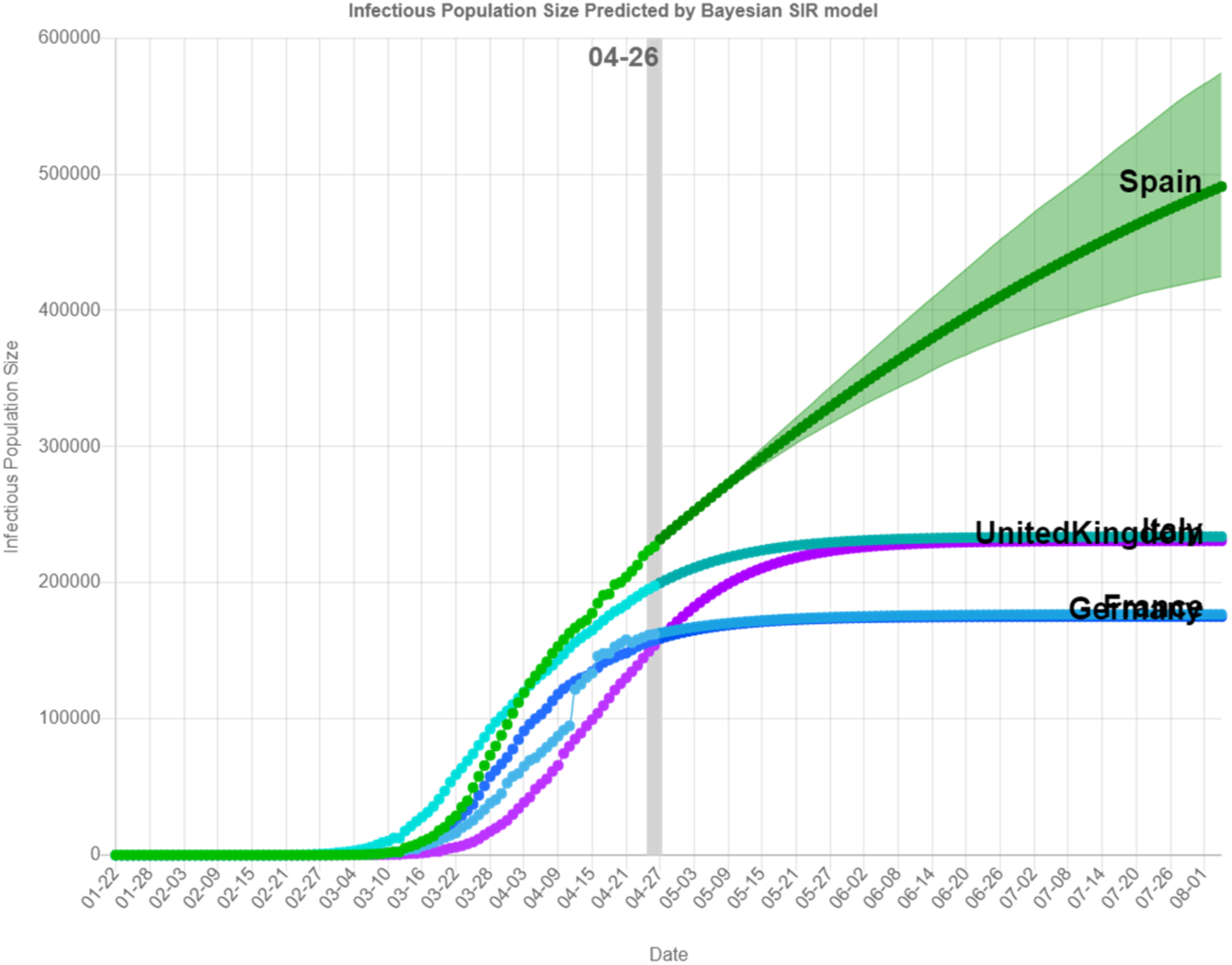
Prediction of the cumulative number of individuals in European countries, estimated with the data up to April 26th.

**Figure 9:**
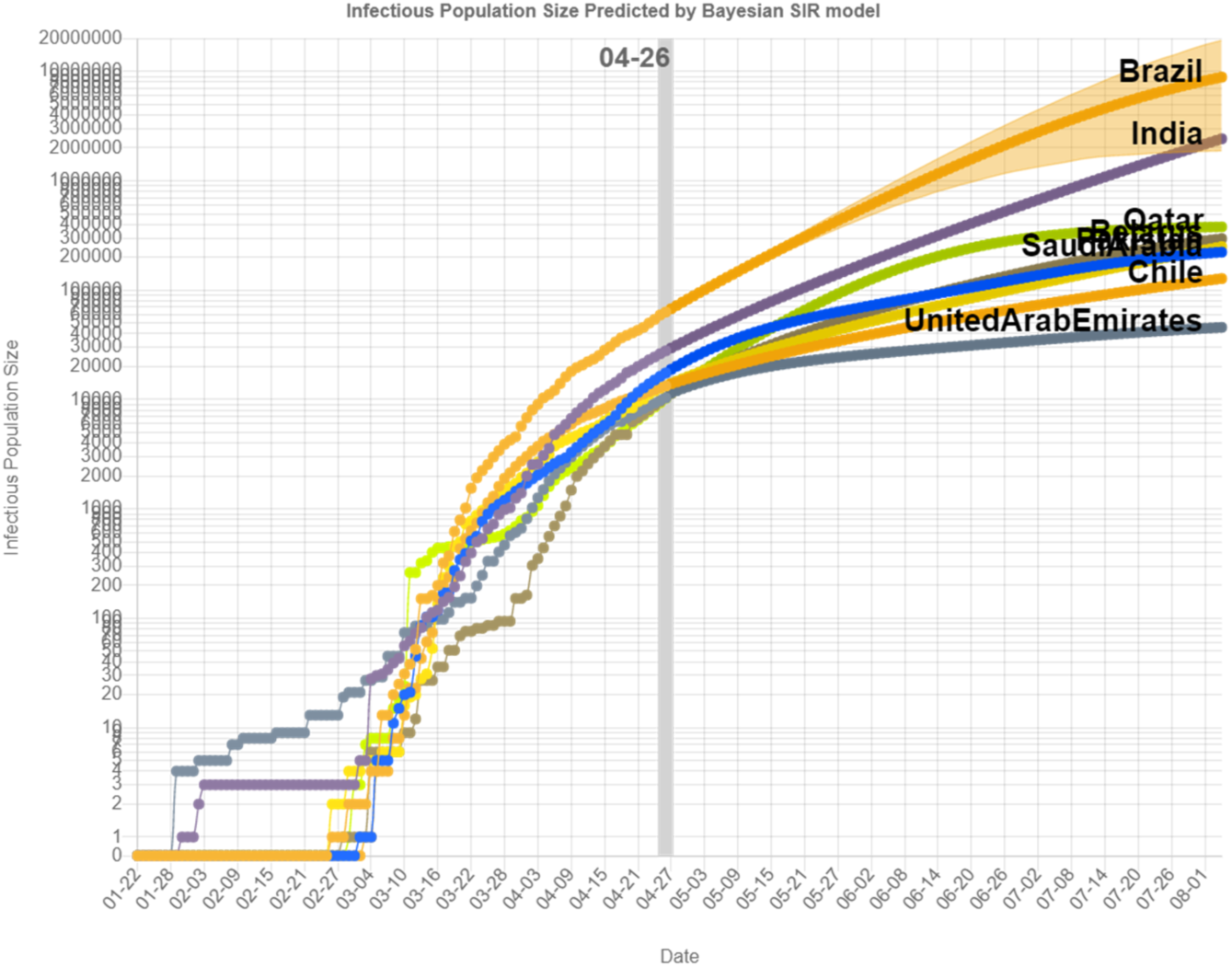
Prediction of the cumulative number of individuals in the countries (Brazil, India, Saudi Arabia, Chile, Pakistan, United Arab Emirates, Belarus, and Qatar, in the descending order at the data analyzed) with predicted increase, estimated with the data up to April 26th.

Next, we also the other countries by particularly focusing on a predicted increasing trend in future. In this analysis, we used the data up to April 26th, and picked up all thirty one countries with 10,000 or more cumulative number of infected individuals (United States with the largest number to Romainia with the smallest number). Most of European countries have already stable state with relatively smaller increasing trend, but Spain and Belrus among the European countries more than 10,000 infectious cases on April 26th, we analyzed. These two countries are still prdicted to show a sharp increasing trend, perhaps these authority needs to be warned with this predictive analysis. Our eyes were catched on the several countries in Central Asia (India and Pakistan), Middle East (Saudi Arabia, United Arab Emirates, and Qatar), and South America (Brazil and Chile). These countries reported relative smaller cumulative number of infected individuals, but our prediction warns the increasing trend in near future is close to exponential. We hope this warning would changed the time course of epidemic dynamics with socio-political action against it.

## 4 Concluding Remark

This research project is on-going and actively improved as long as there is any society under threat of COVID-19, and we believe that the active prediction should publicly contribute to anyone who wishs to consult. Accordingly, we launched the website “Prepidemics” [Hidaka and Torii, 2020], which offers an interactive visualization of the prediction and the predicted results. As a future work, we extend BSIR to mixture model with multiple clusters of system parameters to detect the change of epidemic dynamics due to socio-political action. With this extention, we can assess the effect of social action such as lockdown, more systematically. This kind of quantifation of the social effect would be important to gain and share the effective means against COVID-19.

## Data Availability

This study uses the data repository provided by Johns Hopkins CSSE accessed on April 23rd in 2020.

https://github.com/CSSEGISandData/COVID-19

## Acknowledgements

We are grateful Dr. Miho Fuyama for her discussion on the early version of this manuscript.

